# Post-traumatic Stress Disorder symptom sub-cluster severity predicts gray matter volume changes better than overall symptom severity

**DOI:** 10.1101/2022.07.26.22278078

**Authors:** Sheeva Azma, Rachel Thompson, Diana Bermudez, Rachael Renton, Adebanke Adeyemo, James Meyerhoff, Richard Amdur, Bonnie Green, Mary Ann Dutton, John W. VanMeter

## Abstract

Meta-analysis shows that sub-clusters defined by affected domains of psychosocial functioning capture PTSD subtypes better than symptom clusters defined in the DSM-IV. This pilot study investigated the association between symptom sub-clusters and brain volume in twelve persons with PTSD (females, mean age 40.9 years). Structural magnetic resonance imaging (MRI) images were acquired, and voxel-based morphometry (VBM) was used to estimate local gray matter volume throughout the brain. Participants’ gray matter volume was correlated with both overall PTSD severity and sub-cluster severities. In this preliminary study examining sub-clusters and brain morphometry, we found that neuronal changes associated with sub-clusters may provide a more complete understanding of the neuroanatomical changes that occur in PTSD beyond what can be ascertained using overall disorder severity or comparisons with control subjects. The results of our study suggest that the neurobiological changes resulting from severe trauma depend on the specific sub-clusters of symptoms experienced by individuals with PTSD.

## INTRODUCTION

The neural plasticity resulting from traumatic stress is well characterized (Karatsoreos & McEwen, 2011; McEwen et al., 1999; Weiss, 2007). Lower volume of the hippocampus, medial prefrontal cortex, and insula is characteristic of individuals with Post-Traumatic Stress Disorder (Rauch et al., 2006; Liberzon, 2008). While studies comparing PTSD and non-PTSD individuals have provided essential information about the neurobiological changes induced by severe trauma, the results of these studies tend to be inconsistent (Krause et al., 2007; Landré, 2010; Pacella et al., 2011; Frewen et al., 2011; Hopper et al., 2007).

Individual differences in PTSD symptoms may contribute to the heterogeneity observed in PTSD group difference studies of brain structure (Kroes et al., 2011). Even in the absence of significant correlation between gray matter volume (GMV) and overall PTSD severity, Kroes et al. (2011) found a correspondence between lower GMV and PTSD symptom clusters as defined in the DSM-IV (American Psychiatric Association, 2000). A limitation of Kroes’ study is that the DSM-IV symptom clusters serve to provide physicians with information useful for obtaining a differential diagnosis, rather than providing detail on how specific types of symptoms may be related (e.g. in terms of psychosocial functioning) (Evenden, 1999). Indeed, meta-analysis shows that a four-factor model is preferable to competing models in describing PTSD symptom structure (Yufik and Sims 2010).

The four-factor model, comprised of Hyperarousal, Avoidance, Intrusions, and Dysphoria sub-clusters, demonstrated superior validity and temporal stability in a sample of 396 female survivors of interpersonal violence, 68% of whom were diagnosed with PTSD (Krause et al, 2007).

In the present study, we sought to examine the association between symptom sub-cluster severity, based on the four-factor model, and GMV in patients with PTSD. Understanding the link between sub-clusters and brain volume may illuminate symptom-related biomarkers of PTSD, with implications for treatment.

## METHODS

### Participants

Twelve individuals diagnosed with PTSD related to interpersonal violence (IPV-PTSD) participated in the study. Participants were right-handed females, with mean age of 40.9 years (*SD* = 11.9). All participants experienced a traumatic event since age 18. PTSD diagnosis was assessed using the Clinician-Administered PTSD Scale (CAPS; Blake et al., 1995). All participants completed at least 12 years of education; 9 participants had completed additional college (5 participants, 42%) or graduate/trade school education (4 participants, 33%). Participants were excluded if they endorsed current substance abuse or dependence, lifetime or current psychosis or bipolar disorder, or current suicidal ideation. Individuals taking antipsychotic medications, benzodiazepines, mood stabilizers, clonidine, propanolol, and certain antidepressants such as selective serotonin reuptake inhibitors (SSRIs) or serotonin norepinephrine reuptake inhibitors (SNRIs) were excluded.

Participants completed the PTSD Checklist (PCL; Weathers et al., 1983), a validated self-report measure (Krause et al., 2007), to assess PTSD severity. Participants’ mean score on the PTSD Checklist was 45.5 (*SD* = 9.7), above the recommended cutoff for a diagnosis of PTSD among civilians (Blanchard et al., 1996). Mean subcluster scores for participants are shown in Table 1.

**Table 1.** Overall and four-factor model subcluster scores on the PTSD Checklist (PCL) in all participants, n=12

|  | Mean | Standard deviation |
| --- | --- | --- |
| PCL: Overall | 45.5 | 9.7 |
| PCL: Avoidance subscore | 6.1 | 2.0 |
| PCL: Intrusions subscore | 12.7 | 4.7 |
| PCL: Dysphoria subscore | 21.0 | 7.7 |
| PCL: Hyperarousal subscore | 6.1 | 2.5 |

**Table 2.** Gray Matter Volume (GMV) correlated with total PTSD symptom severity

| Brain region | Cluster-level Corrected p | X (tal) | Y (tal) | Z (tal) | t | Sign of Correlation |
| --- | --- | --- | --- | --- | --- | --- |
| Left Medial Frontal Gyrus | 0.042 | -10.9 | 43.2 | 28.2 | 5.21 | Positive |
| Right Parahippocampal Gyrus (BA28) | 0.049 | 21.8 | 1.7 | -24.5 | -3.25 | Negative |

Participants also completed the Center for Epidemiological Studies Depression Scale (CES-D), a self-report screening questionnaire that measures severity of depressive symptoms (Radloff, 1977). Comorbid depression is present in at least 20% of civilian IPV-PTSD patients (O’Campo et al., 2006). The mean CES-D score in 11 of 12 participants was 42.4 (*SD* = 6.7; one patient had missing data), indicative of a high level of depressive symptoms (Radloff, 1997).

### Procedure

Patients were recruited for the study via ads in several venues. They were screened by telephone for eligibility, then brought to the Clinical Research Center for study procedures. Prior to participating in the experiment, subjects were screened according to MRI safety procedures at Georgetown University. In adhering to Georgetown University Medical Center Institutional Review Board-approved protocols, written consent was obtained following explanation of experimental procedures and each participant was paid $50 for their time.

### Data Acquisition

Magnetic resonance imaging was performed at the Center for Functional and Molecular Imaging at Georgetown University. Structural MRI data was acquired with a 3-Tesla Siemens Trio whole body scanner (Siemens Medical Systems; Erlangen, Germany) with an 8-channel head coil. Brain volumes were acquired using the T1-weighted structural MPRAGE sequence (TR = 1900 ms, TE = 2.52 ms, TI = 900 ms, FOV = 250 × 250 mm^2^, slice thickness = 1 mm, 176 sagittal slices, 246 × 256 matrix).

### Data analysis

Voxel-based morphometry was performed in SPM5 (Ashburner et al., 2000) to estimate local GMV throughout the brain. Preprocessing of the structural MPRAGE volume included spatial normalization to the standard MNI152 template using 12-parameter affine and non-linear cosine basis function transformations, and brain segmentation into gray matter, white matter, and cerebrospinal fluid. Gray matter was then modulated using the Jacobian of the spatial normalization transformation to examine GMV. Spatial smoothing was applied to the modulated GMV with a Gaussian kernel, full-width half-max of 12 mm to reduce noise and to allow for individual variability in the transformation into the MNI atlas space.

To examine the association between PTSD symptoms and GMV, we used SPM5 to investigate correlations between GMV and overall PTSD severity, as well as between GMV and symptom sub-clusters of Intrusions, Dysphoria, Hyperarousal, and Avoidance. Overall PTSD severity was determined by the participant’s score on the PTSD Checklist (PCL; Weathers et al. 1983). Each item on the PCL is included in a unique sub-cluster (Avoidance, Dysphoria, Hyperarousal, or Intrusions). PTSD subcluster scores were calculated for each participant by summing scores for items within a sub-cluster (Krause et al., 2007). We performed multivariate regression analyses with GMV, PCL overall or subcluster score, and age as variables of interest. Effects of participant age on brain structure were controlled for by modelling age as a nuisance regressor. Whole-brain comparisons were performed for each contrast. Statistical maps were thresholded at a whole-brain level *p-value* < 0.005 (t=3.25) and an extent threshold of 10 to form clusters from which final significance was assessed using a cluster-level corrected threshold of *p* < 0.05. In addition, to account for multiple statistical tests conducted for each subcluster, Bonferroni correction was applied to the activation maps of Intrusions, Dysphoria, Hyperarousal, and Avoidance. MNI coordinates of peak activations were transformed to Talairach coordinates using mni2tal (http://imaging.mrc-cbu.cam.ac.uk/downloads/MNI2tal/mni2tal.m).

## RESULTS

Regions of GMV were associated with overall PTSD severity, as reported in Table 1. A positive correlation was observed in left medial frontal gyrus (*p =* 0.042) and a negative correlation was observed in right parahippocampal gyrus (*p* = 0.049). All coordinates are reported in Talairach (“tal”) space.

Several regions of GMV were associated with individual PTSD sub-clusters. Avoidance symptom severity was negatively correlated with GMV in the right insula (BA13; *p* < 0.001), bilateral mid-cingulate gyrus (*p* < 0.001), left inferior temporal gyrus (*p* = 0.009), right superior temporal gyrus (*p* < 0.001), and right middle temporal gyrus (BA 21; *p* < 0.001), and was not positively correlated with GMV in any region of the brain. Hyperarousal symptom severity was positively correlated with GMV in the right anterior cingulate gyrus (BA 32; *p* < 0.001), and was not negatively correlated with GMV in any brain region. There was no significant correlation between severity of Dysphoria symptoms and GMV. Intrusion symptom severity was positively correlated with GMV in the bilateral globus pallidus and putamen (*p* < 0.001). Intrusion symptom severity was negatively correlated with GMV in the bilateral posterior cingulate gyrus (BA 31) and left fusiform gyrus (*p* < 0.001). Table 3 illustrates the brain regions associated with PTSD subcluster severity. All coordinates are reported in Talairach (“tal”) space.

**Table 3.** Gray Matter Volume GMV correlated with PTSD symptom subcluster severity

| *Avoidance* |  |  |  |  |  |  |
| --- | --- | --- | --- | --- | --- | --- |
| Brain region | Cluster-level Corrected p | X (tal) | Y (tal) | Z (tal) | t | Sign of Correlation |
| Right Insula (BA13) | <0.001 | 32.6 | -23.4 | 16.8 | -3.25 | Negative |
| Left Mid Cingulate Gyrus | <0.001 | -9.9 | 7.7 | 36.5 | -3.25 | Negative |
| Right Mid Cingulate Gyrus | <0.001 | 6.9 | 6.6 | 35.6 | -3.25 | Negative |
| Left Inferior Temporal Gyrus | 0.009 | -57.4 | -12.7 | -20.4 | -3.25 | Negative |
| Right Superior Temporal Gyrus | <0.001 | 42.6 | 6.7 | -22.2 | -3.25 | Negative |
| Right Middle Temporal Gyrus (BA 21) | <0.001 | 54.5 | -2.1 | -21.8 | -3.25 | Negative |
| *Hyperarousal* |  |  |  |  |  |  |
| Brain region | Cluster-level Corrected p | X (tal) | Y (tal) | Z (tal) | T | Sign of Correlation |
| Right Anterior Cingulate (BA 32) | <0.001 | 14.9 | 34.2 | 24.1 | 6.23 | Positive |
| *Intrusions* |  |  |  |  |  |  |
| Brain region | Cluster-level Corrected p | X (tal) | Y (tal) | Z (tal) | T | Sign of Correlation |
| Right Globus Pallidus | <0.001 | 12.9 | 2.7 | -3.5 | 5.74 | Positive |
| Right Putamen | <0.001 | 22.8 | -8.5 | 5.0 | 4.44 | Positive |
| Left Putamen | <0.001 | -23.8 | 11.1 | 8.7 | 5.44 | Positive |
| Left Globus Pallidus | <0.001 | -11.9 | 4.9 | 1.6 | 5.15 | Positive |
| Left Posterior Cingulate Gyrus (BA 31) | <0.001 | -12.9 | -43.4 | 25.2 | -3.25 | Negative |
| Right Posterior Cingulate Gyrus (BA 31) | <0.001 | 16.8 | -41.9 | 35.3 | -3.25 | Negative |
| Left Fusiform Gyrus | <0.001 | -46.5 | -14.7 | -22.8 | -3.25 | Negative |

## DISCUSSION

Previous studies have found that overall PTSD severity is associated with decreased GMV in the hippocampus and anterior cingulate cortex, particularly in combat populations. However, studies of non-combat populations (e.g. IPV-PTSD) have reported discrepant results, such as no change in GMV (Landré et al., 2010; Jatzko et al., 2006; Fennema-Notestine et al., 2002; Pederson et al., 2004) or increased GMV in the temporal lobe (Golier et al. 2005). The present study suggests that different symptom sub-cluster profiles may account for the divergent GMV findings observed in patients experiencing various precipitating traumas. Our results reveal a large number of brain areas where GMV is correlated with specific symptom clusters but with overall PTSD severity.

Consistent with a previous study examining DSM-IV-defined symptom clusters (Kroes et al., 2011), we found that severity of sub-clusters were differentially associated with brain alterations. We found that severity of the Avoidance subcluster was associated with decreased GMV in the bilateral middle cingulate gyrus, right insula, and bilateral regions of the temporal lobe. Cingulate dysfunction is a genetically-determined risk factor for PTSD (Pine et al., 2011; Shin et al., 2009) and is associated with the inability to extinguish salient fear associations (Kasai et al., 2008; Quirk et al., 2010). Hopper et al. (2007) reported a negative correlation between activation of the right anterior cingulate and Avoidance symptoms in a script-driven imagery task.

Decreased volume of the temporal cortex has previously been observed in combat-related PTSD. We found that decreased GMV in the superior temporal lobe was associated with heightened Avoidance symptom severity in the present study. In sum, decreased GMV in the cingulate gyrus, insula, and regions of the temporal lobe may be associated with avoidant thoughts and behaviors due to the involvement of these regions in fear extinction, monitoring aversive internal states (Nagai et al., 2007; Strigo et al., 2010), and misattribution of context (Woodward, 2009).

Hyperactive cognitive-appraisal networks in IPV-PTSD may promote hypervigilance symptoms through an exaggerated sensitivity to context cues related to past trauma (Fonzo et al., 2010). Hyperarousal symptoms are mediated by functional interactions between the anterior cingulate cortex and amygdala (Weiss, 2007; Rauch, 2006). We found that hyperarousal symptom severity was positively correlated with GMV in the right anterior cingulate cortex and right inferior parietal lobule. While overall PTSD severity is associated with neuronal loss in the anterior cingulate (Karl et al. 2006; Kasai et al. 2008), such findings may be more pronounced in combat-exposed populations (Landré et al., 2010).

The Intrusions symptom subcluster includes flashbacks, dreams, and reliving of traumatic events. We found that decreased volume in the bilateral posterior cingulate gyrus and left fusiform gyrus, and increased volume in the bilateral globus pallidus and putamen, were associated with Intrusions symptom severity. Consistent with our findings, Lanius et al. (2005) found that PTSD patients with high flashback/reliving symptoms recruited the posterior cingulate cortex over the course of a script-driven imagery fMRI paradigm, whereas control subjects did not. The posterior cingulate plays a role in “assessing the environment and memory” (Vogt et al., 1992); altered function of this region may result in altered self-referential cognition in PTSD (Bluhm, 2012).

Dysphoria symptoms include loss of interest in activities, emotional numbing, and general cognitive disturbance. Despite participants’ high level of severe depressive symptoms, we found no correlation between Dysphoria symptoms and GMV. This is likely due to the restriction of range on the CES-D, where scores were well above the cutoff used to screen for depressive disorders.

To our knowledge, this is the first study investigating the relationship between factor-derived symptom sub-clusters and brain structure in IPV-PTSD. Four-factor sub-clusters, particularly Avoidance, may reflect specific domains of psychosocial functioning (Yufik and Sims 2010; Pietrzak et al., 2010). Patients with IPV-PTSD who are highly dependent on avoidant coping strategies benefit less from therapeutic interventions (Leiner et al., 2012; Pacella et al., 2011) and are at higher risk of escalating symptom severity within the first few months of trauma (Pineles et al., 2011).

Our results extend the findings of Kroes et al. (2011). Despite our smaller sample size and the use of a Bonferroni correction for multiple comparisons, we identified a greater number of brain regions correlating with symptom severity than previously observed by Kroes and colleagues, suggesting that the four-factor sub-cluster model accounts for more neurobiological differences found in PTSD.

In conclusion, our preliminary findings indicate that (1) subcluster-based approaches of investigating gray GMV may reveal neurobiological changes overlooked by traditional between-group studies of PTSD; and (2) neuroimaging studies of symptom sub-clusters may potentially reveal associations between brain structure and psychosocial functioning. Limitations of this pilot study include a small sample size and the large age range of our participants. Future investigations exploring the link between PTSD symptom sub-clusters and trauma type, disease progression, and treatment efficacy may supplement group comparisons of patients and healthy controls in the study of this disorder. In particular, studies that strictly focus on between-group differences may overlook key differences among PTSD subtypes, which this analysis suggests could be an important factor in understanding the neurobiological changes induced by PTSD.

## Data Availability

All data produced in the present study are available upon reasonable request to the authors.

## References

1. American Psychiatric Association. (2000). Diagnostic and statistical manual of mental disorders (4th ed., text rev.) Washington, DC: Author. doi:10.1176/appi.books.9780890423349

2. Ashburner J, Friston KJ. (2000). Voxel-based morphometry—the methods. Neuroimage, 11(6):805–821. doi:10.1006/nimg.2000.0582

3. Blake DD, Weathers FW, Nagy LM, Kaloupek DG, Gusman FD, Charney DS, & Keane TM. (1995). The development of a clinician-administered PTSD scale. Journal of Traumatic Stress, 8, 75–90. doi:10.1007/BF02105408

4. Blanchard EB, Jones-Alexander J, Buckley TC, Forneris CA. (1996). Psychometric properties of the PTSD Checklist (PCL). Behaviour Research and Therapy, 34(8):669–73. doi:10.1016/0005-7967(96)00033-2

5. Bluhm RL, Frewen PA, Coupland NC, Densmore M, Schore AN, Lanius RA. (2012). Neural correlates of self-reflection in post-traumatic stress disorder. Acta psychiatrica Scandinavica, published online 2011 Oct 18. doi:10.1111/j.1600-0447.2011.01773.x.

6. Evenden JL (1999). Varieties of impulsivity. Psychopharmacology, 146:348–361. doi:10.10.1007/PL00005481

7. Fennema-Notestine C, Stein MB, Kennedy CM, Archibald SL, Jernigan TL. (2002). Brain morphometry in female victims of intimate partner violence with and without posttraumatic stress disorder. Biological Psychiatry, 52(11):1089–1101. doi:10.1016/S0006-3223(02)01413-0

8. Fonzo GA, Simmons AN, Thorp SR, Norman SB, Paulus MP, Stein MB. (2010). Exaggerated and disconnected insular-amygdalar blood oxygenation level-dependent response to threat-related emotional faces in women with intimate-partner violence posttraumatic stress disorder. Biological Psychiatry, 68(5):433–41. doi:10.1016/j.biopsych.2010.04.028

9. Frewen PA, Dozois DJ, Neufeld RW, Lane RD, Densmore M, Stevens TK, Lanius RA. (2012). Emotional numbing in posttraumatic stress disorder: a functional magnetic resonance imaging study. Journal of Clinical Psychiatry, 73(4):431–6. doi:10.4088/JCP.10m06477

10. Golier JA, Yehuda R, De Santi S, Segal S, Dolan S, de Leon MJ. (2005). Absence of hippocampal volume differences in survivors of the Nazi Holocaust with and without posttraumatic stress disorder. Psychiatry Research, 139(1):53–64. doi:10.1016/j.pscychresns.2005.02.007

11. Gros DF, Price M, Magruder KM, Frueh BC. (2012). Symptom overlap in posttraumatic stress disorder and major depression. Psychiatry Research, published online 2012 Mar 2. doi:10.1016/j.psychres.2011.10.022

12. Hopper JW, Frewen PA, van der Kolk BA, Lanius RA. (2007). Neural correlates of reexperiencing, avoidance, and dissociation in PTSD: symptom dimensions and emotion regulation in responses to script-driven trauma imagery. Journal of Traumatic Stress, 20(5): 713–25. doi:10.1002/jts.20284

13. Hopper JW, Pitman RK, Su Z, Heyman GM, Lasko NB, Macklin ML, Orr SP, Lukas SE, Elman I. (2008). Probing reward function in posttraumatic stress disorder: expectancy and satisfaction with monetary gains and losses. Journal of Psychiatric Research, 42(10):802–7. doi:10.1016/j.jpsychires.2007.10.008

14. Jatzko A, Rothenhöfer S, Schmitt A, Gaser C, Demirakca T, Weber-Fahr W, Wessa M, Magnotta V, Braus DF. (2006). Hippocampal volume in chronic posttraumatic stress disorder (PTSD): MRI study using two different evaluation methods. Journal of Affective Disorders, 94(1-3):121–126. doi:10.1016/j.jad.2006.03.010

15. Karatsoreos IN, McEwen BS. (2011). Psychobiological allostasis: resistance, resilience, and vulnerability. Trends in Cognitive Science, 15(12):576–84. doi:10.1016/j.tics.2011.10.005

16. Karl A, Schaefer M, Malta LS, Dörfel D, Rohleder N, Werner A. (2006). A meta-analysis of structural brain abnormalities in PTSD. Neuroscience and Biobehavioral Reviews, 30(7):1004–31. doi:10.1016/j.neubiorev.2006.03.004

17. Kasai K, Yamasue H, Gilbertson MW, Shenton ME, Rauch SL, Pitman RK. (2008). Evidence for acquired pregenual anterior cingulate gray matter loss from a twin study of combat-related posttraumatic stress disorder. Biological Psychiatry, 63(6):550–6. doi:10.1016/j.biopsych.2007.06.022

18. Kessler RC, Chiu WT, Demler O, Walters EE. (2005). Prevalence, severity, and comorbidity of twelve-month DSM-IV disorders in the National Comorbidity Survey Replication (NCS-R). Archives of General Psychiatry, 62(6):617–27. doi:10.1001/archpsyc.62.6.617

19. Krause ED, Kaltman S, Goodman L, Dutton MA. (2007). Longitudinal factor structure of posttraumatic stress symptoms related to intimate partner violence. Psychological Assessment, 19(2):165–75. doi:10.1037/1040-3590.19.2.165

20. Kroes MC, Whalley MG, Rugg MD, Brewin CR. (2011). Association between flashbacks and structural brain abnormalities in post-traumatic stress disorder. European Psychiatry, 26(8): 525–31. doi:10.1016/j.eurpsy.2011.03.002

21. Landré L, Destrieux C, Baudry M, Barantin L, Cottier JP, Martineau J, Hommet C, Isingrini M, Belzung C, Gaillard P, Camus V, El Hage W. (2010). Preserved subcortical volumes and cortical thickness in women with sexual abuse-related PTSD. Psychiatry Research, 183(3):181–6. doi:10.1016/j.pscychresns.2010.01.015

22. Lanius RA, Williamson PC, Bluhm RL, Densmore M, Boksman K, Neufeld RW, Gati JS, Menon RS. (2005). Functional connectivity of dissociative responses in posttraumatic stress disorder: a functional magnetic resonance imaging investigation. Biological Psychiatry, 57(8):873–74. doi:10.1016/j.biopsych.2005.01.011

23. Leiner AS, Kearns MC, Jackson JL, Astin MC, Rothbaum BO. (2012). Avoidant coping and treatment outcome in rape-related posttraumatic stress disorder. Journal of Consulting and Clinical Psychology, 80(2):317–21. doi: 10.1037/a0026814

24. Liberzon I, Sripada CS. (2008). The functional neuroanatomy of PTSD: a critical review. Progress in Brain Research, 167: 151–69. doi:10.1016/S0079-6123(07)67011-3

25. McEwen BS, Seeman T. (1999). Protective and damaging effects of mediators of stress. Elaborating and testing the concepts of allostasis and allostatic load. Annals of the New York Academy of Sciences, 896: 30–47. doi:10.1111/j.1749-6632.1999.tb08103.x

26. Nagai M, Kishi K, Kato S. (2007). Insular cortex and neuropsychiatric disorders: a review of recent literature. European Journal of Psychiatry, 22(6):387–94. doi:10.1016/j.eurpsy.2007.02.006

27. O’Campo PO, Kub J, Woods A, Garza M, Jones AS, Gielen AC, Dienemann J, Campbell J. (2006). Depression, PTSD, and comorbidity related to intimate partner violence in civilian and military women. Brief Treatment and Crisis Intervention, 6(2):99–110. doi:10.1093/brief-treatment/mhj010

28. Pacella ML, Irish L, Ostrowski SA, Sledjeski E, Ciesla JA, Fallon W, Spoonster E, Delahanty DL. (2011). Avoidant coping as a mediator between peritraumatic dissociation and posttraumatic stress disorder. Journal of Traumatic Stress, 24(3):317–25. doi:10.1002/jts.20641

29. Pederson CL, Maurer SH, Kaminski PL, Zander KA, Peters CM, Stokes-Crowe LA, Osborn RE. (2004). Hippocampal volume and memory performance in a community-based sample of women with posttraumatic stress disorder secondary to child abuse. Journal of Traumatic Stress, 17(1):37–40. doi:10.1023/B:JOTS.0000014674.84517.46

30. Pietrzak RH, Goldstein MB, Malley JC, Rivers AJ, Southwick SM. (2010). Structure of posttraumatic stress disorder symptoms and psychosocial functioning in Veterans of Operations Enduring Freedom and Iraqi Freedom. Psychiatry Research, 178(2):323–9. doi:10.1016/j.psychres.2010.04.039

31. Pine DS, Freedman R. (2011). Imaging a brighter future. American Journal of Psychiatry, 168(9):885–7. doi:10.1176/appi.ajp.2011.11050788

32. Pineles SL, Mostoufi SM, Ready CB, Street AE, Griffin MG, Resick PA. (2011). Trauma reactivity, avoidant coping, and PTSD symptoms: a moderating relationship? Journal of Abnormal Psychology, 120(1):240–6. doi:10.1037/a0022123

33. Quirk GJ, Milad MR. (2010). Neuroscience: Editing out fear. Nature, 463(7277):36–7. doi:10.1038/463036a

34. Radloff LS (1977). The CES-D scale: A self-report depression scale for research in the general population. Applied Psychological Measurement, 1, 385–401. doi: doi:10.1177/014662167700100306

35. Rauch, S.L., Shin, L.M., Phelps, E.A. (2006). Neurocircuitry models of posttraumatic stress disorder and extinction: human neuroimaging research – past, present, and future. Biological Psychiatry, 60(4):376–82. doi:10.1016/j.biopsych.2006.06.004

36. Shin LM, Lasko NB, Macklin ML, Karpf RD, Milad MR, Orr SP, Goetz JM, Fischman AJ, Rauch SL, Pitman RK. (2009). Resting metabolic activity in the cingulate cortex and vulnerability to posttraumatic stress disorder. Archives of General Psychiatry, 66(10):1099–107. doi:10.1001/archgenpsychiatry.2009.138

37. Strigo IA, Simmons AN, Matthews SC, Grimes EM, Allard CB, Reinhardt LE, Paulus MP, Stein MB. (2010). Neural correlates of altered pain response in women with posttraumatic stress disorder from intimate partner violence. Biological Psychiatry, 68(5):442–50. doi:10.1016/j.biopsych.2010.03.034

38. Vogt BA, Finch DM, Olson CR. (1992). Functional heterogeneity in cingulate cortex: the anterior executive and posterior evaluative regions. Cerebral Cortex, 2(6):435–43. doi:10.1093/cercor/2.6.435-a

39. Weathers F, Litz B, Herman D, Huska J, Keane T. (1993). The PTSD Checklist (PCL): Reliability, Validity, and Diagnostic Utility. Annual Meeting of International Society for Traumatic Stress Studies, San Antonio, TX. doi:10.1016/0005-7967(96)00033-2

40. Weiss SJ. (2007). Neurobiological alterations associated with traumatic stress. Perspectives in Psychiatric Care, 43(3): 114–22. doi:10.1111/j.1744-6163.2007.00120.x

41. Woodward SH, Schaer M, Kaloupek DG, Cediel L, Eliez S. (2009). Smaller global and regional cortical volume in combat-related posttraumatic stress disorder. Archives of General Psychiatry, 66(12): 1373–82. doi:10.1001/archgenpsychiatry.2009.160

42. Yufik T, Simms LJ. (2010). A meta-analytic investigation of the structure of posttraumatic stress disorder symptoms. Journal of Abnormal Psychology, 119(4): 764–776. doi:10.1037/a0020981

